# Hospital admission with non-alcoholic fatty liver disease is associated with increased all-cause mortality independent of cardiovascular risk factors

**DOI:** 10.1101/2019.12.24.19015750

**Authors:** Jake P. Mann, Paul Carter, Matthew J. Armstrong, Hesham K Abdelaziz, Hardeep Uppal, Billal Patel, Suresh Chandran, Ranjit More, Philip N. Newsome, Rahul Potluri

**Author notes:** **Corresponding author:** Dr. Jake P Mann, MRC Epidemiology Unit, Level 3, Institute of Metabolic Science, Addenbrooke’s Hospital, Cambridge, CB2 0QQ. These authors contributed equally to this work (Philip N. Newsome, Rahul Potluri). **Author contributions: JPM:** Conceptualization, formal analysis, Writing – Original Draft Preparation, Writing – Review & Editing. **PC:** Data curation, formal analysis, Writing – Review & Editing. **MA:** Conceptualization, Writing – Review & Editing. **HKA, HU, BP, SC, RM:** Data Curation, Project Administration, Resources, Writing – Review & Editing. **PNN & RP:** Conceptualization, Project Administration, Supervision, Writing – Review & Editing.

## Abstract

**Background:** Non-alcoholic fatty liver disease (NAFLD) is common and strongly associated with the metabolic syndrome. Though NAFLD may progress to end-stage liver disease, the top cause of mortality in NAFLD is cardiovascular disease (CVD). Most of the data on liver-related mortality in NAFLD derives from specialist liver centres. We aimed to assess mortality in NAFLD when adjusting for CVD in a ‘real world’ cohort of inpatients.

**Methods:** Retrospective study of hospitalised patients with 14-years follow-up. NAFL (non-alcoholi c fatty liver), non-alcoholic steatohepatitis (NASH), and NAFLD-cirrhosis groups were defined by ICD-10 codes using ACALM methodology. Cases were age-/sex-matched 1:10 with non-NAFLD hospitalised patients from the ACALM registry. All-cause mortality was compared between groups using cox regression adjusted for CVD and metabolic syndrome risk factors.

**Results:** We identified 1238 patients with NAFL, 105 with NASH and 1235 with NAFLD-cirrhosis. There was an increasing burden of cardiovascular disease with progression from NAFL to NASH to cirrhosis. After adjustment for demographics, metabolic syndrome components and cardiovascular disease, patients with NAFL, NASH, and cirrhosis all had increased all-cause mortality (HR 1.3 (CI 1.1-1.5), HR 1.5 (CI 1.0-2.3) and HR 3.5 (CI 3.3-3.9), respectively). Hepatic decompensation (NAFL HR 8.0 (CI 6.1-10.4), NASH HR 6.5 (2.7-15.4) and cirrhosis HR 85.8 (CI 72-104)), and hepatocellular carcinoma were increased in all NAFLD groups.

**Conclusion:** There is a high burden of cardiovascular disease in NAFLD-cirrhosis patients. From a large “real-life” non-specialist registry of hospitalized patients, NAFLD patients have increased overall mortality and rate of liver-related complications compared to controls after adjusting for cardiovascular disease.

## Introduction

Non-alcoholic fatty liver disease is the most common liver disease in Europe[1] and is strongly associated with all features of the metabolic syndrome[2]. The majority of NAFLD patients have simple steatosis (non-alcoholic fatty liver, NAFL) and only a minority with non-alcoholic steatohepatitis (NASH), with or without fibrosis. However a small, but significant proportion do progress to end-stage liver disease[3].

NAFLD is thought to be associated with increased all-cause and cardiovascular mortality[4– 6]. It has been established that fibrosis is the main predictor of long-term liver-related morbidity in NAFLD[7–9] and that patients with NASH but no fibrosis have a similar outcome to those with NAFL and no fibrosis. However, these studies included biopsy-proven patients in specialist clinics and therefore there is likely significant ascertainment bias in estimating rates of hepatic complications. The natural history of NAFLD and its impact upon clinical services is an important topic that divides expert opinion[10,11].

Cardiovascular disease (CVD) is the commonest cause of mortality in patients with NAFLD[9]. A recent large-scale analysis strongly suggests that this is due to prevalence of classical CVD risk factors such as type 2 diabetes and dyslipidaemia[12]. Insulin resistance is thought to be the primary driver linking all these features of the metabolic syndrome. In response to the positive energy balance of obesity, subcutaneous adipose becomes dysfunctional and there is expansion of visceral white adipocytes, which are less insuli n sensitive and have a higher basal rate of lipolysis[13]. Elevated insulin and increased substrate delivery to the liver promotes hepatic steatosis by driving increased *de novo* lipogenesis without increasing glucose uptake[14]. Cumulatively, this results in a rise in circulating triglycerides, impaired low-density lipoprotein clearance, and higher serum glucose. Hepatic steatosis is also thought to alter the composition of secreted lipoparticles [15].

A further increasingly important consideration is the burden of cardiovascular co-morbidity in patients with end-stage NAFLD for whom transplantation is an option[16]. There is currently limited data on the prevalence of CVD in patients with NAFLD cirrhosis[17]. CVD events are common post-transplant sequelae and chronic kidney disease is linked to reduced graft survival[18].

Whilst several previous natural history studies have included comparison to age- and gender-matched control populations, they have been unable to control for CVD[19–21]. Therefore, it remains unclear whether NAFLD is associated with increased all-cause mortality after correction for cardiac and metabolic disease risk factors.

We aimed to first describe the burden of cardiovascular disease across the NAFLD disease spectrum: non-alcoholic fatty liver (NAFL, or simple steatosis), non-alcoholic steatohepatitis (NASH, steatosis plus histological inflammation), and NAFLD-cirrhosis (end-stage fibrosis) and whether NAFL and NASH are associated with increased all-cause mortality in a real life cohort of hospitalised UK patients from the ACALM registry, after correction for CVD and metabolic risk factors.

## Materials and Methods

The study was conducted as a retrospective cohort study of adult patients in England during 2000-2013 who were admitted to 7 different hospitals with naturalistic follow-up. All available data was included. Tracing of anonymised patients was performed using the ACALM (Algorithm for Comorbidities, Associations, Length of stay and Mortality) study protocol to develop the ACALM registry and has been previously described by our group[22–27]. Briefly, medical records were obtained from local health authority computerized Hospital Activity Analysis register, which is routinely collected by all NHS hospitals. This provides fully anonymized data on hospital admissions and allows for the long-term tracing of patients at an individual hospital. The ACALM protocol uses using International Classification of Disease, 10^th^ edition (ICD-10) and Office of Population Censuses and Surveys Classification of Interventions and Procedures (OPCS-4) coding systems to trace patients. This data was obtained separately for the seven included hospitals. Similar data could be obtained through national Hospital Episode Statistics or from any local Hospital Activity Analysis register.

ICD-10 codes were used to identify patients with NAFL (non-alcoholic fatty liver, K76.0), NASH (non-alcoholic steatohepatitis, K75.8), and NAFLD-cirrhosis (cryptogenic cirrhosis, K74.6). Patients with a history of alcohol excess (F10) were excluded. Where a patient was coded with both NAFL and NASH, they were included in the NASH group. Patients coded with both NAFL and NAFLD-cirrhosis, or NASH and NAFLD-cirrhosis, were included in the cirrhosis group. As per UK practice, the diagnosis of NAFL, NASH, or NAFLD-cirrhosis were made according to clinical judgement and the latest guidelines but the results of the investigations used to derive the diagnoses were not available. An age- and sex-matched control group (with no liver-related diagnoses) was identified from the same ACALM registry and matched 10:1 to patients with NAFLD diagnoses.

All of these patients were then assessed for the presence of several cardiovascular co-morbidities and risk factors, including: congestive heart failure (CHF, I150.0), atrial fibrillation (I48), and non-insulin dependent diabetes mellitus (NIDDM, E11), chronic kidney disease (N18), obesity (E66.0), myocardial infarction (I21-I22), ischaemic heart disease (I20-25), ischaemic stroke (I63.9), hyperlipidaemia (E78.5), hypertension (I10), and peripheral vascular disease (I73.9). Patients were also assessed for liver-related events: hepatocellular carcinoma (C22.9), hepatic failure (K72), oesophageal varices (I85), portal hypertension (K76.6), splenomegaly (R16.1), and ascites (R18). A combined ‘hepatic decompensation or failure’ score was generated from the sum of all non-malignant liver-related events. Inclusi on of hepatic encephalopathy (K72.9) or variceal bleeding (I98.3, I98.8, I85.9) did not identify any additional patients. Jaundice was not included due to identification of patients with obstructive (non-hepatic) jaundice.

Vital status (alive or deceased) on 31^st^ March 2013 was determined by record linkage to the National Health Tracing Services (NHS strategic tracing service) and was received along with the raw data; this was used to calculate all-cause mortality and survival.

The first admission to hospital treatment was chosen as index admission, follow-up of patients continued until 31^st^ March 2013. Confidentiality of information was maintained in accordance with the UK Data Protection Act. The patient data included was fully anonymous and non-identifiable when received by the authors, and collected routinely by the hospitals. Therefore, according to local research ethics policies we were not required to seek formal ethical approval for this study.

Data analysis was performed using SPSS version 20.0 (SPSS Inc. Chicago, IL) and GraphPad Prism version 7.0. Clinical outcomes were compared between groups using chi-squared tests. Cox regression analysis was used to determine adjusted hazard ratios for overall mortality in NAFL, NASH, and cirrhosis groups relative to control. Cox regression was performed twice, first accounting for variations in demographics (age, gender, ethnicity), and then accounting for demographics plus obesity, NIDDM, CHF, ischaemic stroke, myocardial infarction, chronic kidney disease, peripheral vascular disease, hypertension, hyperlipidaemia, ischaemic heart disease, and atrial fibrillation. Participants with incomplete data were excluded. Kaplan-Meier curves were used to determine survival in patients. Multivariate logistic regression was used to determine hazard ratios for hepatic failure/decompensation and hepatocellular carcinoma, adjusted for demographics (age, gender, ethnicity). P<0.05 was taken as significant. No additional sensitivity analyses were performed.

## Results

2,578 patients were identified with NAFLD-spectrum diagnoses, of which 1,238 had NAFL, 105 had NASH and 1,235 NAFLD-cirrhosis (Table 1). They were matched to 25,780 hospitalised in-patient controls. The median duration of follow-up for each group was: control 5.3 years, NAFL 4.6 years, NASH 4.4 years, and cirrhosis 2.8 years (range 1 day - 14 years for all).

**Table 1.** Demographics, mortality, and liver outcomes for control, NAFL, NASH and NAFLD-cirrhosis patients.

| **Clinical outcome**<br>**Number (%)** | **Control**<br>**(n=25,780)** | **NAFL**<br>**(n=1,238)** | **NASH**<br>**(n=105)** | **Cirrhosis**<br>**(n=1,235)** |
| --- | --- | --- | --- | --- |
| Mean age (±standard error) | 55 ±15 | 51 ±14 | 52 ±17 | 59 ±14 |
| Male | 14,570<br>(56.5%) | 690<br>(55.7%) | 57 (54.3%) | 710 (57.5%) |
| Caucasian | 20,626<br>(80.0%) | 960<br>(77.5%) | 87 (82.9%) | 943 (76.4%) |
| South Asian | 1,693 (6.6%) | 150<br>(12.1%) | 6 (5.7%) | 151 (12.2%) |
| All-cause mortality | 3,852<br>(14.9%) | 174<br>(14.1%) | 23 (21.9%) | 664 (53.8%) |
| Overall mortality adjusted for<br>demographic characteristics<br>(HR with 95% CI) | - | 1.4 (1.2-1.6) | 2.0 (1.3-3.0) | 4.1 (3.7-4.4) |
| Overall mortality adjusted for demographics, CV risk factors, and CV disease (HR with 95% CI) | - | 1.3 (1.1-1.5) | 1.5 (1.0-2.3) | 3.5 (3.3-3.9) |
| Hepatic failure/decompensation, n (%) | 232 (0.9%) | 85 (6.9%) | 6 (5.7%) | 551 (44.6%) |
| Adj. HR (95% CI) | - | 8.0 (6.1-10.4) | 6.5 (2.7-15.4) | 85.8 (72-102) |
| Hepatocellular carcinoma, n (%) | 49 (0.2%) | 14 (1.1%) | 1 (1.0%) | 152 (12.3%) |
| Adj. HR (95% CI) | - | 3.8 (2.0-7.5) | 2.6 (0.3-21.0) | 65.4 (45-94) |

Clinical outcomes for patients during a 14-year study period. Adjusted hazard ratios of overall mortality for NAFL, NASH, and cirrhosis are relative to control. Adjustment for demographic characteristics included gender, age and ethnicity. Adjustment for cardiovascular risk factors and cardiovascular disease includes: obesity, type 2 diabetes mellitus, CHF, ischaemic stroke, myocardial infarction, chronic kidney disease, peripheral vascular disease, hypertension, hyperlipidaemia, ischaemic heart disease, and atrial fibrillation. Adjusted hazard ratios for liver outcomes are corrected for demographic characteristics. Adj., adjusted; CI = confidence interval; CV = cardiovascular; HR = hazard ratio.

Patients with NAFL and NASH had a higher prevalence of metabolic risk factors and cardiovascular disease than hospitalised controls (Table 2). Hyperlipidaemia, type 2 diabetes mellitus (T2DM), and obesity were similar between NAFL and NASH groups. Compared to the NAFL group, patients with cirrhosis were more likely to have T2DM (20% NAFL vs. 30% cirrhosis, p<0.001) but had a lower prevalence of hyperlipidaemia (12% NAFL vs. 4.8% cirrhosis, p<0.001).

**Table 2.** Cardiovascular disease burden across the NAFLD spectrum.

| **Clinical outcome** | **Control** | **NAFL** | **NASH** | **Cirrhosis** |
| --- | --- | --- | --- | --- |
| **Number (%)** | **(n=25,780)** | **(n=1238)** | **(n=105)** | **(n=1235)** |
| Obesity | 318 (1.2) | 89 (7.2)<sup>##</sup> | 11 (10.5) | 41 (3.3)<sup>***</sup> |
| Type 2 Diabetes | 2,414 (9.4) | 246 (19.9)<sup>##</sup> | 21 (21.9) | 372 (30.1)<sup>***</sup> |
| Hyperlipidaemia | 2,065 (8.0) | 149 (12.0)<sup>##</sup> | 19 (18.1) | 59 (4.8)<sup>***</sup> |
| Congestive Heart Failure | 897 (3.5) | 46 (3.7) | 11 (10.5)<sup>‡</sup> | 81 (6.6)<sup>**</sup> |
| Atrial fibrillation | 1,212 (4.7) | 61 (4.9) | 9 (8.6) | 102 (8.3)<sup>***</sup> |
| Chronic Kidney Disease | 522 (2.0) | 33 (2.7) | 5 (4.8) | 81 (6.6)<sup>***</sup> |
| Ischaemic heart disease | 3,027 (11.7) | 121 (9.8) | 11 (10.5) | 152 (12.3)<sup>*</sup> |
| Myocardial infarction | 966 (3.7) | 24 (1.9) | 4 (3.8) | 27 (2.2) |
| Ischaemic stroke | 521 (2.0) | 34 (2.5) | 0 | 34 (2.8) |
| Hypertension | 5,808 (22.5) | 365 (29.5)<sup>##</sup> | 37 (35.2) | 333 (27.0) |
| Peripheral vascular disease | 393 (1.5) | 13 (1.1) | 3 (2.9) | 16 (1.3) |
| **All-cause mortality** | **3,841 (14.9)** | **175 (14.1)** | **23 (21.9)<sup>†</sup>** | **664 (53.8)<sup>\*\*\*</sup>** |

Crude rates of mortality, metabolic, and cardiovascular outcomes for NAFL, NASH, and cirrhosis, patients during a 14-year study period.
For control vs. NAFL: <sup>#</sup> $p<0.05$ , <sup>##</sup> $p<0.001$ For NAFL vs. NASH: <sup>†</sup> $p<0.05$ , <sup>‡</sup> $p<0.001$ . For NAFL vs. cirrhosis: <sup>\*</sup> $p<0.05$ , <sup>\*\*</sup> $p<0.001$ , <sup>\*\*\*</sup> $p<0.0001$ . p values were calculated using chi squared tests.

There was an increasing burden of cardiovascular co-morbidity with more advanced liver disease. Compared to the control group, patients with NAFL had increased hypertension (30% NAFL vs. 23% control, p<0.001.) Patients with NASH had a higher prevalence of heart failure than those with NAFL (11% vs. 4%, p=0.001). The cirrhosis group showed higher prevalence of heart failure, atrial fibrillation, CKD, and ischaemic heart disease, compared to the NAFL group.

Unadjusted 14-year all-cause mortality was 14.9% for patients in the control group, 14.1% for patients with NAFL, 21.9% for patients with NASH and 53.8% for those with NAFLD-cirrhosis (Fig 1). After adjustment for age, gender and ethnicity, all-cause mortality hazard ratios (HR) were higher in all NAFLD groups compared to the control group (Table 1). After adjustment for cardiovascular factors all-cause mortality was still elevated compared to the control group: NAFL HR 1.3 (CI 1.1-1.5), NASH HR 1.5 (1.0-2.3) and NAFLD-cirrhosis HR 3.5 (CI 3.3-3.9). All cause-mortality was similar between NAFL and NASH groups.

**Figure 1.**
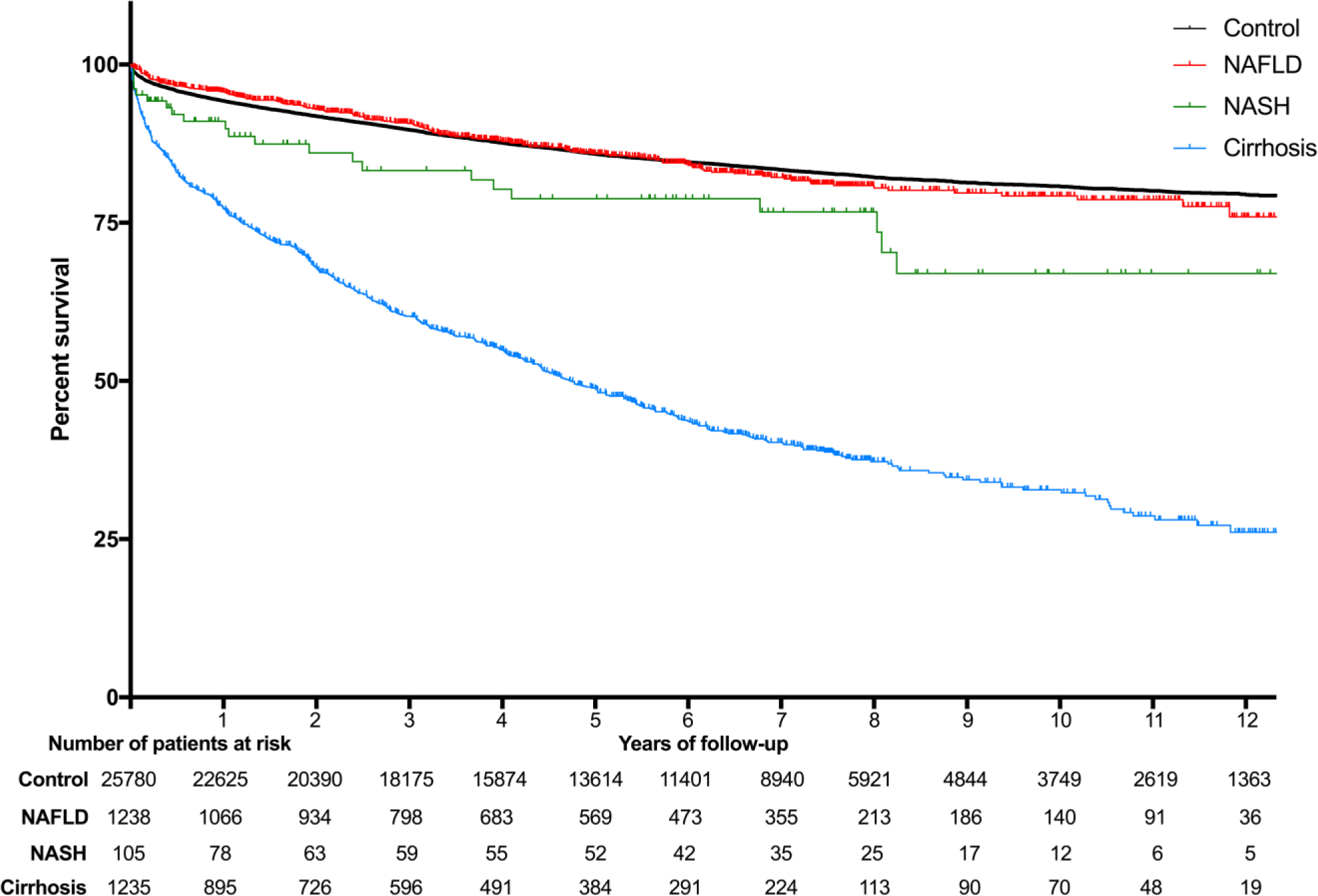
Kaplan-Meier survival curves for NAFL, NASH, and cirrhosis patients compared to non-NAFLD patients admitted to hospital.

The prevalence of hepatic events (hepatic failure and development of HCC) was higher in all groups of patients with NAFLD and was similar between NAFL and NASH groups. This observation remained after correction for age, sex, and ethnicity: for hepatic failure or decompensation, relative to control, NAFL HR 8.0 (CI 6.1-10.4), NASH HR 6.5 (2.7-15.4) and NAFLD-cirrhosis HR 85.8 (CI 72-104, Table 1); and for HCC, relative to control, NAFL HR 3.8 (2.0-7.5), NASH HR 2.6 (0.3-21.0), NAFLD-cirrhosis HR 65.4 (45-94).

## Discussion

This study provides important non-specialist “real life” data amongst hospitalised patients demonstrating an increased mortality for patients with NAFLD, irrespective of fibrosis, even after adjustment for CVD. The size of the cohort and lack of link to specialist liver centres reduces likelihood of bias. These data will help inform healthcare demand for this cohort of patients, complementing modelling estimates[28,29]. In addition, we have highlighted the burden of cardiovascular disease in patients with NAFLD-cirrhosis, which highlights a particular issue for transplantation. Whilst all participants in our cohort were hospitalised, which may increase their risk of future clinical events, so were the controls, thus the comparisons between groups remain valid.

The strong association between NAFLD and cardiovascular disease has been well established[30,31]. Relationships have been identified between NAFLD and heart failure[32], atrial fibrillation[33], hypertension[34], stroke[35], chronic kidney disease[36], and coronary artery disease. NAFLD has even been linked to increased mortality in acute heart failure[37]. However strong observational data from a large European meta-analysis suggests that NAFLD is not causal in acceleration of cardiovascular disease[12]. Our data highlights the particularly increased prevalence towards cirrhosis. Indeed, hypertension has recently been highlighted as an independent risk factor for advanced fibrosis in NAFLD. A further consideration is whether heart failure contributes to accelerated fibrosis in NAFLD[38], though causality is difficult to establish.

Previous studies with biopsy-defined cohorts have been smaller, did not adjust for cardiovascular diagnoses[9,20], and found no difference in mortality between participants with NAFLD and no fibrosis and controls. Kim *et al*. used NHANES data to stratify patients by non-invasive fibrosis scores, and again found no increase in mortality in patients with ultrasound-defined NAFLD, after correction for diabetes and hypertension[19]. This may be accounted for by differences in the ethnicity of the cohort and also the general, rather than specialist, nature of our population.

NAFLD may itself be a marker of sub-clinical cardiovascular disease. For example, increased carotid intima media thickness has been found in adolescents with NAFLD[39]. This is mechanistically plausible as hepatic steatosis occurs (in part) in response to peripheral insulin resistance and elevated substrate delivery from lipolysis of adipose tissue. Steatosis itself then contributes to systemic insulin resistance[40]. Therefore, in this analysis, despite adjusting for metabolic covariates and cardiovascular risk factors, elevated mortality may reflect the sub-clinical nature of atherosclerosis associated with NAFLD, even at an early stage.

Given the shared disease mechanisms and clinical outcomes for NAFLD and cardiovascular disease, these data suggest a common framework for treatment. Weight loss is the only established treatment strategy for NAFLD[41] and there is data suggesting that specific dietary regimens (including the Mediterranean diet) are beneficial[42]. The same lifestyle interventions and aggressive risk factor modification will dual impact on reducing cardiovascular[43,44] and hepatic events.

Whilst we were not able to determine cause of death or admission in our cohort, we were able to determine that liver decompensation events were increased in all groups relative to the control group.This study is limited by its retrospective design and the use of generic coding, which did not provide information on how the diagnosis was obtained i.e. imaging, liver function tests, or liver biopsy and we were unable to identify whether NAFLD was the cause of admission or an existing co-morbidity in cases. Therefore, differences between the NAFL and NASH groups should be interpreted with this in mind. However, coding improvements in recent years along with standardised diagnosis of NAFLD in the UK means that the impact of inaccurate coding is likely to be low. Published studies suggest that only 2-10%[1] of the NAFLD groups may be affected and thus have minimal impact on the results observed in this study. The use of ICD-10 codes for the exclusion of other causes of liver dysfuncti on (for example, viral hepatitis) and patients with a history of alcohol consumption may have also contributed to inaccurate coding. However, such biases have been limited from our previous study looking at the association between cardiovascular and respiratory conditions [30]. There is likely to be a degree of under coding of NAFLD, especially as clinical awareness of NAFLD was not optimal at the beginning of the data capture[47].

In conclusion, these results contribute to our understanding of co-morbidity, mortality and liver decompensation in patients with NAFLD spectrum disease and demonstrate that the increase in mortality occurs independently of known cardiovascular risk factors.

## Data Availability

All summary statistics available at request

## Abbreviations

ACALM: Algorithm for Comorbidities, Associations, Length of stay and Mortality
CV: cardiovascular
HR: hazard ratio
ICD-10: International Classification of Disease, 10^th^ edition
NAFLD: non-alcoholic fatty liver (disease)
NASH: non-alcoholi c steatohepatitis

## References

1 Younossi ZM, Koenig AB, Abdelatif D, Fazel Y, Henry L, Wymer M. Global epidemiology of nonalcoholic fatty liver disease—Meta-analytic assessment of prevalence, incidence, and outcomes. Hepatology. 2016;64: 73–84. doi:10.1002/hep.28431

2 Stefan N, Häring H-U, Cusi K. Non-alcoholic fatty liver disease: causes, diagnosis, cardiometabolic consequences, and treatment strategies. Lancet Diabetes Endocrinol. 2018;8587: 1–12. doi:10.1016/S2213-8587(18)30154-2

3 Angulo P, Kleiner DE, Dam-Larsen S, Adams L a, Bjornsson ES, Charatcharoenwitthaya P, et al. Liver Fibrosis, but No Other Histologic Features, Is Associated With Long-term Outcomes of Patients With Nonalcoholic Fatty Liver Disease. Gastroenterology. 2015;149: 389–397.e10. doi:10.1053/j.gastro.2015.04.043

4 Musso G, Gambino R, Maurizio C, Pagano G. Meta-analysis: Natural history of non-alcoholic fatty liver disease (NAFLD) and diagnostic accuracy of non-invasive tests for liver disease severity. Ann Med. 2011;43: 617–649.

5 Hamaguchi M, Kojima T, Takeda N, Nagata C, Takeda J, Sarui H, et al. Nonalcolholic fatty liver disease is a novel predictor cardiovascular disease. World J Gastroenterol. 2007;13: 1579–1584. doi:10.3748/wjg.v13.i10.1579

6 Wild SH, Walker JJ, Morling JR, McAllister DA, Colhoun HM, Farran B, et al. Cardiovascular disease, cancer, and mortality among people with type 2 diabetes and alcoholic or nonalcoholic fatty liver disease hospital admission. Diabetes Care. 2018;41: 341–347. doi:10.2337/dc17-1590

7 Hagström H, Nasr P, Ekstedt M, Hammar U, Stål P, Hultcrantz R, et al. Fibrosis stage but not NASH predicts mortality and time to development of severe liver disease in biopsy-proven NAFLD. J Hepatol. 2017; doi: 10.1016/j.jhep.2017.07.027. doi:10.1016/j.jhep.2017.07.027

8 Dulai PS, Singh S, Patel J, Soni M, Prokop LJ, Younossi Z, et al. Increased risk of mortality by fibrosis stage in nonalcoholic fatty liver disease: Systematic review and meta-analysis. Hepatology. 2017;65: 1557–1565. doi:10.1002/hep.29085

9 Angulo P, Kleiner DE, Dam-Larsen S, Adams LA, Bjornsson ES, Charatcharoenwitthaya P, et al. Liver fibrosis, but no other histologic features, is associated with long-term outcomes of patients with nonalcoholic fatty liver disease. Gastroenterology. 2015;149: 389–397.e10. doi:10.1053/j.gastro.2015.04.043

10 Rowe IA. Too much medicine: overdiagnosis and overtreatment of non-alcoholic fatty liver disease. Lancet Gastroenterol Hepatol. 2018;3: 66–72. doi:10.1016/S2468-1253(17)30142-5

11 Ratziu V. The painful reality of end-stage liver disease in NASH. Lancet Gastroenterol Hepatol. 2018;3: 8–10.

12 Alexander M, Loomis AK, van der Lei J, Duarte-Salles T, Prieto-Alhambra D, Ansell D, et al. Non-alcoholic fatty liver disease and risk of incident acute myocardial infarction and stroke: findings from matched cohort study of 18 million European adults. Bmj. 2019;367: 5367. doi:10.1136/bmj.l5367

13 Samuel VT, Shulman GI. Nonalcoholic Fatty Liver Disease as a Nexus of Metabolic and Hepatic Diseases. Cell Metab. 2017. doi:10.1016/j.cmet.2017.08.002

14 Sanders FWB, Acharjee A, Walker C, Marney L, Roberts LD, Imamura F, et al. Hepatic steatosis risk is partly driven by increased de novo lipogenesis following carbohydrate consumption. Genome Biol. 2018;19: 79. doi:10.1186/s13059-018-1439-8

15 Anstee QM, Targher G, Day CP. Progression of NAFLD to diabetes mellitus, cardiovascular disease or cirrhosis. Nat Rev Gastroenterol Hepatol. 2013;10: 330–344. doi:10.1038/nrgastro.2013.41

16 Younossi ZM, Marchesini G, Pinto-Cortez H, Petta S. Epidemiology of Nonalcoholic Fatty Liver Disease and Nonalcoholic Steatohepatitis: Implications for Liver Transplantation. Transplantation. 2018; doi: 10.1097/TP.0000000000002484 [Epub ahead of pr. doi:10.1097/TP.0000000000000000

17 Kilaru SM, Quarta G, Popov V. Diabetes, Heart Failure, and Chronic Kidney Disease are Risk Factors for Nafld-Related Cirrhosis: A Nationwide Analysis. Gastroenterology. 2017;152: S688–S689. doi:10.1016/S0016-5085(17)32415-0

18 Molnar MZ, Joglekar K, Jiang Y, Cholankeril G, Abdul MKM, Kedia S, et al. Association of Pre-Transplant Renal Function with Liver Graft and Patient Survival after Liver Transplantation in Patients with Nonalcoholic Steatohepatitis. Liver Transplant. 2018; doi: 10.1002/lt.25367 [Epub ahead of print]. doi:10.1002/lt.25367

19 Kim D, Kim WR, Kim HJ, Therneau TM. Association between noninvasive fibrosis markers and mortality among adults with nonalcoholic fatty liver disease in the United States. Hepatology. 2013;57: 1357–1365. doi:10.1002/hep.26156

20 Ekstedt M, Hagström H, Nasr P, Fredrikson M, Stal P, Kechagias S, et al. Fibrosis stage is the strongest predictor for disease-specific mortality in NAFLD after up to 33 years of follow-up. Hepatology. 2015;61: 1547–1554. Available: http://scholar.google.com/scholar?hl=en&btnG=Search&q=intitle:No+Title#0

21 Ekstedt M, Franzén LE, Mathiesen UL, Thorelius L, Holmqvist M, Bodemar G, et al. Long-term follow-up of patients with NAFLD and elevated liver enzymes. Hepatology. 2006;44: 865–73. doi:10.1002/hep.21327

22 Sangha J, Natalwala A, Mann J, Uppal H, Mummadi SM, Haque A, et al. Co-morbidities and mortality associated with intracranial bleeds and ischaemic stroke. Int J Neurosci. 2015;125: 256–263. doi:10.3109/00207454.2014.930463

23 Uppal H, Chandran S, Potluri R. Risk factors for mortality in Down syndrome. J Intellect Disabil Res. 2015; [Epub ahead of print]. doi:10.1111/jir.12196

24 Potluri R, Baig M, Mavi JS, Ali N, Aziz A, Uppal H, et al. The role of angioplasty in patients with acute coronary syndrome and previous coronary artery bypass grafting. Int J Cardiol. 2014;176: 760–3. doi:10.1016/j.ijcard.2014.07.097

25 Carter P, Rai G, Aziz A, Mann J, Chandran S, Uppal H, et al. Trends of cardiovascular disease amongst psychiatric patients between 2001 and 2012 in Greater Manchester, UK. Int J Cardiol. 2014;173: 573–574.

26 Ziff OJ, Carter PR, McGowan J, Uppal H, Chandran S, Russell S, et al. The interplay between atrial fibrillation and heart failure on long-term mortality and length of stay: Insights from the, United Kingdom ACALM registry. Int J Cardiol. 2018;252: 117–121. doi:10.1016/j.ijcard.2017.06.033

27 Ziaei F, Zaman M, Rasoul D, Gorantla RS, Bhayani R, Shakir S, et al. The prevalence of atrial fibrillation amongst heart failure patients increases with age. Int J Cardiol. 2016;214: 410–411. doi:10.1016/j.ijcard.2016.03.198

28 Younossi ZM, Blissett D, Blissett R, Henry L, Stepanova M, Younossi Y, et al. The economic and clinical burden of nonalcoholic fatty liver disease in the United States and Europe. Hepatology. 2016;64: 1577–1586. doi:10.1002/hep.28785

29 Estes C, Razavi H, Loomba R, Younossi ZM, Sanyal AJ. Modeling the epidemic of nonalcoholic fatty liver disease demonstrates an exponential increase in burden of disease. Hepatology. 2017; doi:10.1002/hep.29466. doi:10.1002/hep.

30 Wu S, Wu F, Yingying D, Hou J, Bi J, Zhang Z. Association of non-alcoholic fatty liver disease with major adverse cardiovascular events: A systematic review and meta-analysis. Sci Rep. 2016;6: 33386. doi:10.1038/srep33386

31 Hagstrom H, Nasr P, Ekstedt M, Hammar U, Stal P, Askling J, et al. Cardiovascular risk factors in non-alcoholic fatty liver disease. Liver Int. 2018; 1–8. doi:10.1111/liv.13973

32 Wijarnpreecha K, Lou S, Panjawatanan P, Cheungpasitporn W, Pungpapong S, Lukens FJ, et al. Association between diastolic cardiac dysfunction and nonalcoholic fatty liver disease: A systematic review and meta-analysis. Dig Liver Dis. 2018;50: 1166–1175. doi:10.1016/j.dld.2018.09.004

33 Targher G, Valbusa F, Bonapace S, Bertolini L, Zenari L, Rodella S, et al. Non-Alcoholic Fatty Liver Disease Is Associated with an Increased Incidence of Atrial Fibrillation in Patients with Type 2 Diabetes. PLoS One. 2013;8. doi:10.1371/journal.pone.0057183

34 Sung KC, Wild SH, Byrne CD. Development of new fatty liver, or resolution of existing fatty liver, over five years of follow-up, and risk of incident hypertension. J Hepatol. 2014;60: 1040–1045. doi:10.1016/j.jhep.2014.01.009

35 Alexander KS, Zakai NA, Lidofsky SD, Callas PW, Judd SE, Tracy RP, et al. Non-alcoholic fatty liver disease, liver biomarkers and stroke risk: The reasons for geographic and racial differences in stroke cohort. PLoS One. 2018;13: 1–13. doi:10.1371/journal.pone.0194153

36 Paik J, Golabi P, Younoszai Z, Mishra A, Trimble G, Younossi ZM. Chronic Kidney Disease is Independently Associated with Increased Mortality in Patients with Nonalcoholic Fatty Liver Disease. Liver Int. 2018; doi: 10.1111/liv.13992 [Epub ahead of print]. doi:10.1111/liv.13992

37 Valbusa F, Agnoletti D, Scala L, Grillo C, Arduini P, Bonapace S, et al. Non-alcoholic fatty liver disease and increased risk of all-cause mortality in elderly patients admitted for acute heart failure. Int J Cardiol. 2018;265: 162–168. doi:10.1016/j.ijcard.2018.04.129

38 Colli A, Pozzoni P, Berzuini A, Gerosa A, Canovi C, Molteni EE, et al. Decompensated Chronic Heart Failure: Increased Liver Stiffness Measured by Means of Transient Elastography. Radiology. 2010;257: 872–878. doi:10.1148/radiol.10100013

39 Gökçe S, Atbinici Z, Aycan Z, Cınar HG, Zorlu P. The relationship between pediatric nonalcoholic fatty liver disease and cardiovascular risk factors and increased risk of atherosclerosis in obese children. Pediatr Cardiol. 2013;34: 308–15. doi:10.1007/s00246-012-0447-9

40 Samuel VT, Shulman GI. The pathogenesis of insulin resistance: Integrating signaling pathways and substrate flux. J Clin Invest. 2016;126: 12–22. doi:10.1172/JCI77812

41 Dixon JB, Bhathal PS, Hughes NR, O’Brien PE. Nonalcoholic fatty liver disease: Improvement in liver histological analysis with weight loss. Hepatology. 2004;39: 1647–1654. doi:10.1002/hep.20251

42 Properzi C, O’Sullivan TA, Sherriff JL, Ching HL, Jeffrey GP, Buckley RF, et al. Ad libitum Mediterranean and Low Fat Diets both Significantly Reduce Hepatic Steatosis: a Randomized Controlled Trial. Hepatology. 2018;68: 1741–1754. doi:10.1002/hep.30076

43 Widmer RJ, Flammer AJ, Lerman LO, Lerman A. The Mediterranean diet, its components, and cardiovascular disease. Am J Med. 2015;128: 229–238. doi:10.1016/j.amjmed.2014.10.014

44 Poirier P, Giles TD, Bray GA, Hong Y, Stern JS, Pi-Sunyer FX, et al. Obesity and cardiovascular disease: Pathophysiology, evaluation, and effect of weight loss: An update of the 1997 American Heart Association Scientific Statement on obesity and heart disease from the Obesity Committee of the Council on Nutrition, Physical. Circulation. 2006;113: 898–918. doi:10.1161/CIRCULATIONAHA.106.171016

45 Zeb I, Li D, Budoff MJ, Katz R, Lloyd-Jones D, Agatston A, et al. Nonalcoholic Fatty Liver Disease and Incident Cardiac Events the Multi-Ethnic Study of Atherosclerosis. J Am Coll Cardiol. 2016;67: 1965–1966. doi:10.1016/j.jacc.2016.01.070

46 Targher G, Byrne CD, Lonardo A, Zoppini G, Barbui C. Non-alcoholic fatty liver disease and risk of incident cardiovascular disease: A meta-analysis. J Hepatol. 2016;65: 589–600. doi:10.1016/j.jhep.2016.05.013

47 Sherwood P, Lyburn I, Brown S, Ryder S. How are abnormal results for liver function tests dealt with in primary care? Audit of yeild and impact. BMJ. 2001;322: 276–278.

